# Predicting responses to chemotherapy from nutrition in triple negative breast cancer patients (The PRE-NUTRITIVE Study): protocol for a prospective feasibility study

**DOI:** 10.1101/2025.03.21.25324389

**Authors:** Giorgia Cioccoloni, Ibtihal Barnawi, Amy Burkinshaw, Sue Hartup, Alex Rzeszutek, Nisha Sharma, Elton J. R. Vasconcelos, Stephen D. Hursting, Thomas A Hughes, Baek Kim, James L. Thorne

## Abstract

**Background:** Adherence to the World Cancer Research Fund (WCRF) and American Institute for Cancer Research (AICR) cancer prevention guidelines is linked to lower cancer incidence and improved outcomes. However, the relationship between these guidelines and chemotherapy response, particularly in Triple Negative Breast Cancer, is not well understood. TNBC has the poorest survival rates among breast cancer subtypes, with only 32% of patients achieving pathological complete response after neoadjuvant chemotherapy. Predicting which patients will respond and gain survival benefits remains a challenge and identifying patients unlikely to respond would help provide more effective treatment options and reduce side-effects and hospital admissions. This study assesses the feasibility of collecting data for a clinical trial aimed at identifying factors that predict chemoresponse with particular attention on diet, nutrition, physical activity, adherence to WCRF/AICR recommendations, and tumour and circulating biomarkers.

**Methods:** This prospective, non-randomised feasibility study will recruit, over 24 months, between 15-20 triple negative breast cancer patients undergoing neoadjuvant chemotherapy. The data collected are: body mass index, chemotherapy details, surgery type, gene expression analysis in diagnostic tumour cores, serum and plasma samples for lipid and vitamin analysis, tumour response by magnetic resonance imaging during and after treatment and pathological response after treatment. Participants will complete patient-reported outcome measures, food and physical activity questionnaires, at the start and end of treatment.

**Discussion:** This study aims to explore the impact of dietary patterns on chemotherapy responses in TNBC patients, a subtype with poor prognosis and high relapse risk. Adherence to the WCRF/AICR cancer prevention guidelines is linked to reduced cancer incidence and better outcomes. However, the role of diet in predicting chemotherapy response remains unclear. The study seeks to gather data for a future clinical trial examining these connections, aligning with research priorities to prevent cancer relapse and provide evidence-based dietary advice. This feasibility study will inform patient recruitment, data collection, and trial design.

**Trial registration:** This trial was prospectively registered on 12^th^ December 2022 (ISRCTN20130557).

## BACKGROUND

Adherence to the World Cancer Research Fund (WCRF)/American Institute for Cancer Research (AICR) cancer prevention guidelines is associated with reduced risk of cancer [1, 2] and improved outcomes for cancer survivors [3]. Overcoming chemoresistance is a significant challenge in cancer treatment and optimising therapy efficacy remains an unmet challenge, especially in difficult to cure cancers that are still treated with systemic cytotoxic chemotherapy. Clinical tools which can predict the most effective chemotherapy regimen for an individual patient remain undeveloped, but if available would aid in clinical decision making in determining the optimal treatment regimen to improve survival outcomes and would reducing hospital visits and side-effects.

The roles of nutrient signalling, metabolism, and dietary patterns are yet to be systematically explored in the context of chemotherapy efficacy. However, several examples show that modification of these pathways alter accumulation of toxic intermediates and end-products of metabolism and impact on processing of pharmaceuticals [4, 5]. For example citrus is contraindicated for ~30% of all prescription drugs owing to irreversible inhibition of the detoxification enzyme CYP3A4[6]; PPAR ligands such as vitamin B3 induce CYP2C8, a phase 2 drug detoxification factor [7]; vitamin A derivatives change expression of phase 1 drug efflux pumps such as P-glycoprotein that are linked to chemoresistance in several cancer types [8–10]; cholesterol metabolites induce chemotherapy resistance via P-glycoprotein in triple negative breast cancer (TNBC) [11]; vitamin D [12–14] and curcumin [15] increase drug efficacy.

Triple negative breast cancers (TNBC) lack expression of oestrogen (ER), progesterone (PR), and HER2 receptors. This renders conventional targeted therapies ineffective, leaving chemotherapy, and more recently immunotherapy, as the mainstay systemic therapeutic options for TNBC patients. Survival in TNBC remains the lowest of all breast cancer subtypes and accounts for approximately 10–15% of breast cancer cases in White patients [16] but can be up to 65% of cases in Black populations [17–20] underlining a large racial disparity in incidence and outcomes for this cancer of unmet clinical need. Typically, TNBC patients have surgery to excise the tumour followed by adjuvant treatments including radiotherapy and adjuvant chemotherapy (ACT) to eliminate potential residual microscopic disease. Increasingly, patients are recommended neo-adjuvant chemotherapy (NACT) to downstage the tumour and thus reduce the surgical burden. NACT also provides an opportunity to monitor tumour response to therapy via interval MRI scans and on resection pathology (residual cancer burden). If a sub-optimal tumour response is observed, the chemotherapy regimen and choice of cytotoxic agents can be altered. Despite this, the proportion of TNBC patients achieving pathological complete response (pCR) with NACT is 32% [21]. Predicting which TNBC patients are less likely to respond optimally to NACT would enable opportunity to tailor more effective chemotherapy regimen and offer other existing or novel targeted treatment with an aim of improving patient survival.

Despite adherence to WCRF/AICR recommendations being associated with improved outcomes in several cancer types, the interaction between the molecular changes driven by guideline adherence and response to chemotherapy in TNBC patients has not been explored. This study will explore the feasibility of collecting the data required for a clinical trial that will aim to identify dietary, nutritional, and physical activity parameters, including adherence to the WCRF/AICR cancer prevention recommendations, which may influence and/or determine chemotherapy response in TNBC.

## METHODS

### Study aims

This is a feasibility study that will collect data on the nutritional status, dietary patterns, WCRF/AICR adherence, physical activity of TNBC patients, the intra-tumoral expression of nutritional and metabolic related genes, blood lipids, and chemotherapy efficacy. PRE-NUTRITIVE trial will assess the feasibility of collecting this information in the clinical setting, recruitment rate, and identify barriers to data collection and analysis, and provide data for power calculations for a subsequent clinical trial. The secondary aim is to collect preliminary data that may indicate involvement of specific nutrient/metabolic parameters that may predict chemoresponse in TNBC patients, although the study is underpowered to achieve this secondary aim.

### Study objectives

1. Assess the feasibility of collecting nutritional metrics from TNBC patients during standard clinical care pathways.
2. Assess feasibility of collecting tumour biopsy, blood samples and tumour response to chemotherapy data matched to patient’s nutritional data.
3. Establish patient recruitment rate.
4. Assess whether adherence to WCRF/AICR cancer prevention recommendations can be determined.
5. Assess active metabolic pathways.
6. Test power of candidate metabolic pathways to predict chemoresponse and inform power calculations for a future study.
7. Establish feasibility of designing a sufficiently powered multicentre clinical trial to develop a chemoresponse prediction tool.

### Study design

The PRE-NUTRITIVE study is a prospective, non-randomised, feasibility, cohort study. A minimum of fifteen and maximum of twenty TNBC patients undergoing NACT at Leeds Teaching Hospitals NHS Trust (LTHT) will be recruited over 24 months and followed through their treatment. After consent, pseudonymised information relevant to the study will be collected from the hospital electronic patient record (EPR), such as: body mass index (BMI), type/frequency/duration of chemotherapy, tumour response to NACT using MRI, type of breast and axillary surgery performed, pathology assessment of diagnostic biopsy and surgical resection. Prior to starting NACT, two tumour core biopsies will be collected during marker during clip placement, or at an extra visit (not mandatory for participation), and analysed by RNA-sequencing to characterise gene expression patterns. Study participants will complete patient reported outcome measures (PROMs), a validated food-frequency questionnaire (EPIC-Norfolk [22, 23]), food diary (myfood24 [24]), and survey of their physical activity levels (EPAQ2 [25, 26]) at the start of NACT to evaluate baseline adherence to the WCRF recommendations, and at the end of NACT to evaluate any potential dietary and physical activity changes. Study participants will also be asked to provide a blood sample at the start and end of NACT to ascertain blood lipid and micro- and macro-nutrient levels in plasma/serum. Tumour tissue will be collected during surgical resection (under surgeon’s decision) and compared to pre-treatment biopsy at RNA level. In addition, this will allow the analysis of tumour material from patients who were unable to provide biopsy due to prior marker clip placement or other reasons.

### Study settings

This trial will be run jointly at Leeds Teaching Hospitals Trust (LTHT) and School of Food Science and Nutrition at University of Leeds. Patient recruitment, along with all the previously cited activities on study participants, will be performed at LTHT. The School of Food Science and Nutrition at University of Leeds will perform all sample and data analyses.

### Eligibility criteria

#### Inclusion criteria

Consenting participants must be:

1. Female or male patients aged 18 years or older
2. Uni- or bilateral TNBC in the breast and/or axilla (including primary, second primary, locoregional recurrence, or inflammatory breast cancer)
3. Clinical recommendation for NACT
4. Ability to provide informed consent
5. Uni- or multi-focal invasive breast cancer of any histological subtype (if multifocal, dominant lesion that is triple negative to be biopsied) and any tumour (T) and clinical node (N) staging.

#### Exclusion criteria

1. Patients diagnosed with pure non-invasive breast cancer (*e*.*g*. DCIS) or with non-TNBC invasive subtypes
2. Patients diagnosed with distant metastatic disease at the time of primary breast cancer diagnosis
3. Inability to provide informed written consent

TNM staging system has been stated as per Cancer Research UK definitions[27].

### Consent

After identification of potential participants, the direct clinical team and an appropriately trained member of the hospital team will provide a patient information sheet (PIS). Patients will be given a minimum of 24 hours to consider the study and enable sufficient time to consider the information and ask any questions they may have about participation. For patients who agree to participate in the study, they will sign and date the latest approved version of the informed consent form (ICF) enabling the study specific procedures and data collection to proceed. Written consent can be taken by the direct clinical team and/or research nurses. To reduce burden, completion of the consent form will occur during the patient’s scheduled hospital visits. In addition, patients will have the additional option of undergoing postal consent (*e*.*g*. due to changes in COVID-19 restrictions). Participants undergoing face-to-face or postal consent will be given a fully signed copy of the ICF before study participation. For the ancillary/future studies, only data and samples of study participants who provide this additional optional consent to reuse their data/sample will be used.

### Outcomes

#### Primary feasibility outcomes

Determine feasibility of evaluating patient’s nutritional status via PROMs, nutritional profiling of blood, and collecting tumour biopsy.

#### Secondary exploratory outcomes

1. Patient uptake rate of blood and tumour tissue donation
2. Completion rate of patient survey and barriers to completion
3. Measurement of variation in nutritional status and adherence between study participants (based on molecular assays and questionnaires)
4. Accuracy and compliance of WCRF guideline adherence using food-frequency questionnaire (FFQ), myfood24 and physical activity questionnaire-based methods of data collection
5. RNA yield from tumour biopsies and selection of best candidate metabolic pathways that predict chemoresponse.
6. Measurement of predictors of chemoresponse by gene expression analysis.

### Participant timeline

The participant timeline and schedule of the intervention is outlined in Table 1 and Figure 1.

**Table 1.** Participant timeline from initial screening to end of study.

|  | STUDY PERIOD |  |  |  |  |  |
| --- | --- | --- | --- | --- | --- | --- |
|  | Enrolment | Post-enrolment |  |  |  | Close-out |
| TIMEPOINT | $-t_0$ | $t_0$ | $t_1$ | $t_2$ | $t_3$ | $t_x$ |
| **ENROLMENT** |  |  |  |  |  |  |
| Eligibility screen | X |  |  |  |  |  |
| Informed consent | X |  |  |  |  |  |
| **DATA/SAMPLE COLLECTION AND PROCESSING** |  |  |  |  |  |  |
| Blood samples collection and Micro-/macronutrient blood quantification |  | X |  |  | X |  |
| Biopsy samples collection and Tumour RNA analysis |  | X |  |  | X |  |
| FFQ (EPIC-Norfolk) |  | X |  |  | X |  |
| EPAQ2 |  | X |  |  | X |  |
| Myfood24 |  | X |  |  | X |  |
| Breastfeeding and smoking data |  | X |  |  |  |  |
| Magnetic Resonance Imaging (MRI) data |  | X | X | X |  |  |
| Anthropomorphic measures (BMI and waist-hip circumference) |  | X |  |  |  |  |
| Residual cancer burden data |  |  |  |  | X |  |
| Pathological assessment data |  |  |  |  | X |  |

| ASSESSMENT |  |  |  |  |  |  |
| --- | --- | --- | --- | --- | --- | --- |
| Feasibility of nutritional metrics collection |  |  |  |  |  | X |
| Recruitment rate |  |  |  |  |  | X |
| Intra-tumor variability |  |  |  |  |  | X |
| Feasibility multicenter trial for prediction model |  |  |  |  |  | X |

\*\*t0: baseline, t1: second NACT cycle; t2: last NACT cycle; t3: surgery. Body Mass Index (BMI); EPIC Physical Activity Questionnaire 2 (EPAQ2), European Prospective Investigation into Cancer (EPIC), food-frequency questionnaire (FFQ), Magnetic Resonance Imaging (MRI), Neoadjuvant Chemotherapy (NACT).

**Figure 1.**
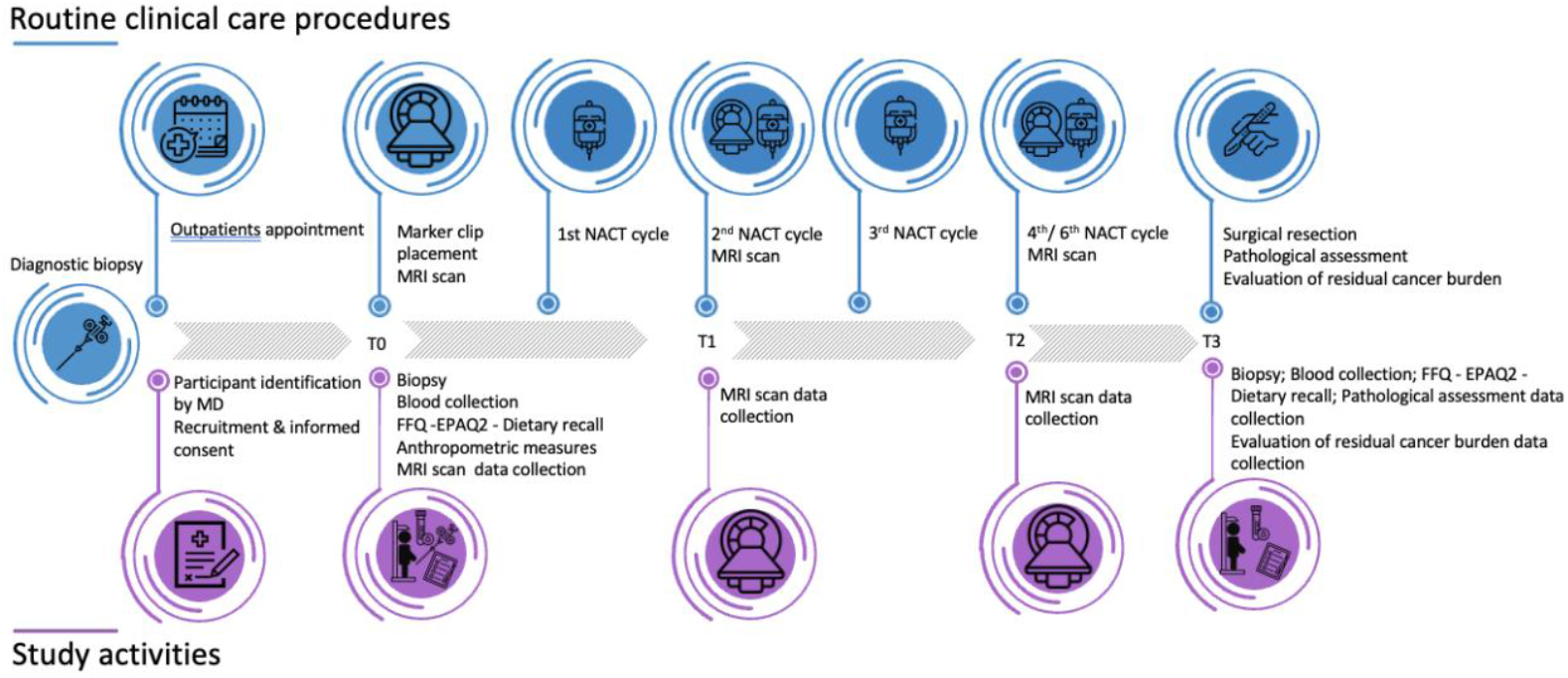
The PRE-NUTRITIVE Study participant timeline Timeline for study ‘participants. In blue, routine clinical care procedures. In purple, PRE-NUTRITIVE Study activities. EPIC Physical Activity Questionnaire 2 (EPAQ2), European Prospective Investigation into Cancer (EPIC), food-frequency questionnaire (FFQ), medical doctor (MD), Magnetic Resonance Imaging (MRI), Neoadjuvant Chemotherapy (NACT). Figure was created with the use of Slidesgo and Freepik (https://slidesgo.com/; https://www.freepik.com/).

### Recruitment

Patient recruitment, along with all the previously cited activities on study participants, will be performed by patient’s direct clinical team at LTHT. Study observation period starts from when the study participants sign the informed consent and continues until the time when the patient finishes NACT and completes surgical treatment and pathology assessment.

### Data collection, management and analysis

#### Data collection methods

Participant will undergo anthropometric measurements and tumour biopsy collection at baseline (T0). Tumour tissue will also be collected during surgery at the end of the study (T3). At baseline and at tumour surgical resection (T3) participants will donate fasted blood samples and complete surveys. Data from Magnetic Resonance Imaging (MRI) scans will be collected at baseline, and at concurrently to the second and last NACT cycle (T1, and T2 respectively). Pathological assessment and evaluation of residual cancer burden data will be collected after tumour resection (T3).

#### Blood samples collection

Blood collection (up to 50mL) will be performed in fasting state (>8 hours fast). The samples will be labelled and immediately transported to the School of Food Science and Nutrition at University of Leeds laboratories to isolate serum/plasma. Serum/plasma samples will be then stored before being analysed using validated scientific methods to quantify micro-/macronutrient content.

#### Biopsies for RNA analysis

The patient will be biopsied under US guidance with local anaesthesia where 2 × 14G core biopsies will be obtained from the dominant mass by a member of the radiology team trained to perform biopsies. Similarly, 2 × 14G core biopsies will be obtained from the dominant mass by the trained surgical team once the cancer has been resected from the patient. The samples will be labelled, placed in RNA later on ice, and immediately transported to the School of Food Science and Nutrition at University of Leeds laboratories to be processed for RNA-sequencing.

#### Dietary recording and assessment of adherence to the WCRF/AICR cancer prevention recommendations

*Patient Reported Outcome Measures (PROMs)* Validated FFQ (EPIC-Norfolk) for UK population [22, 23] and myfood24 [24] food diary will be conducted to record dietary intake. Adherence to six 2018 WCRF/AICR recommendations about healthy diet (Eat wholegrains, vegetables, fruit and beans; limit ‘fast foods’; limit red and processed meat; limit sugar sweetened drinks; limit alcohol consumption; do not use supplements for cancer prevention)[28]. Adherence to the WCRF/AICR recommendations will be scored according to previous studies Winkels *et. al*., [29], Romaguera *et. al*., [30], Malcomson *et. al*., [31].

Physical activity EPAQ2 survey [25, 26] will be conducted to determine adherence to 2018 WCRF/AICR recommendations about physical activity. Information on current or previous breastfeeding and smoking/tobacco use will be collected to determine adherence to 2018 WCRF/AICR recommendations[28].

#### Magnetic Resonance Imaging (MRI)

MRI scan will be performed at baseline prior to commencing NACT as standard of care. Briefly, MRI is performed at 1.5 T with the patient lying prone in a bilateral breast coil. T1 and T2-weighted volume imaging of both breasts, pre-contrast, will be performed.

#### Anthropomorphic measures

Body weight will be measured to the nearest 0.1 kg, using a technical balance and height with a stadiometer to the nearest 0.1 cm. Body weight and height will be used to calculate the body mass index (BMI) (BMI=weight (kg)/height (m)^2^) of each subject to assess the adherence to the WCRF/AICR recommendations regarding body fatness. Waist and hip circumferences will be measured with a tape measure to the nearest 0.5 cm.

#### Pathological assessment and evaluation of residual cancer burden

After surgical resection of tumour, specimen pathological assessment will be performed as per standard of care with recording of residual cancer[32].

#### Data management

Data collection will occur in accordance with good clinical practice (GCP), Caldicott principles and the General Data Protection Regulation (GDPR) 2018 and will work in line with NHS confidentiality guidelines and codes of conduct. Data for each patient will be pseudonymised using a unique alphanumeric study identification number. Personal data will only be available to the research team at the LTHT. Codes will be created for each participant personal information and data pseudonymised. Information will be held securely on paper and electronically at LTHT and University of Leeds. Direct access of coded data will be granted to the research team at the University of Leeds, authorised representatives from the Sponsor and the regulatory authorities to permit trial-related monitoring, audits and inspections in line with participant consent. Fully anonymized research data will be deposited with the University of Leeds data repository. Raw sequencing data will be stored by LeedsOmics facility for 5 years and then deposited and hosted with a digital object identifier (DOI).

### Monitoring

The members of the Trial Management Group (TMG) (JLT, SH, EJRV) are responsible for the day-to-day management of the trial and will oversee all aspects of the conduct of the trial. The Sponsor will monitor and audit the conduct of research as required, periodically reviewing safety data and issues. The site TMG member and PI (SH) will be responsible for data recording, CRF, serious adverse effects (SAEs), reports, notifications, applications and submissions availability and that these documents are accurate, complete, dated and identify the trial. Any medical condition present at baseline where the severity and/or frequency do not get worse during the study and NACT-related toxicity is not considered as adverse effect (AE). Only SAEs related and unexpected occurring to a research participant will be recorded and reported to the main REC and to the Sponsor.

### Protocol amendments

The CI (JLT) and the Sponsor’s authorised representative will be responsible for the decision to amend the protocol and for deciding whether an amendment is substantial or non-substantial. Relevant extracts or new versions of revised documents will be submitted to the REC, showing the new version number and date and giving both the previous and new wording which is clearly identifiable. Amendments will be tracked in the protocol appendix and the version of the protocol will be updated. To date, four amendments were made, submitted and approved. This protocol paper reflects the most recent protocol version (version 1.4, 04.04.2024).

### Dissemination plans

On completion of the trial, the data will be analysed and tabulated and a Final Trial Report prepared to be sent to REC-HRA the within 12 months of the end of the study. Data will be published in scientific journals, which will depend on the outcome of data analysis. University of Leeds will curate and deposit data in the European Nucleotide Archive with a unique DOI. Overall study outcomes will also be made freely available to trial participants. Full trial report, pseudonymised participant level dataset, and statistical code for generating the results will not be publicly available.

## Discussion

Adherence to the cancer prevention guidelines established by the WCRF/AICR consistently associates with reduced cancer incidence and improved cancer outcomes across diverse cancer types[2, 3, 31, 33, 34]. Notably, the exploration of nutrient signalling, metabolism, and dietary patterns in relation to chemotherapy efficacy remains as a significant knowledge gap in predicting chemotherapy non-responders. The PRE-NUTRITIVE study addresses this by focusing specifically on the impact of dietary patterns on chemotherapy responses in patients with TNBC, a cancer subtype characterized by poor prognosis and heightened risk of relapse[21].

Our investigation aligns with broader research priorities outlined by the National Cancer Research Institute (NCRI) and James Lind Alliance[35], specifically by aiming to identify effective ways to prevent cancer relapse and support cancer patients in lifestyle and health by providing evidence-based robust dietary advice. Utilizing the WCRF/AICR cancer prevention guidelines as a foundational framework, our study aims to explore the feasibility of collecting comprehensive data for a subsequent clinical trial that will explore the connections between dietary patterns and chemotherapy response through observation and targeted nutritional interventions.

The information obtained during the PRE-NUTRITIVE feasibility study will provide key information on data collection and processing, recruitment, and patient engagement that is required for a large prospective, randomised trial, powered to detect whether differences in adherence to the WCRF cancer prevention guidance could influence chemotherapy efficacy. Furthermore, data collected during the PRE-NUTRITIVE study will facilitate robust power calculations, establishing the groundwork for a multicentre clinical trial with sufficient statistical power to investigate the relationship between dietary patterns and chemotherapy response.

In conclusion, this trial aims to extend understanding of the interplay between dietary factors and chemotherapy response and lays the foundation for future work that aims to improve outcomes for TNBC and ultimately other cancer patients being treated with systemic chemotherapy.

## Data Availability

This is a trial design protocol and no data are contained in this manuscript

## List of abbreviations

ACT: Adjuvant Chemotherapy
AE: Adverse Event
mAICR: American Institute for Cancer Research
AR: Adverse Reaction
BMI: Body Mass Index
CRF: Case Report Form
CRO: Contract Research Organisation
CTA: Clinical Trial Authorisation
EPAQ2: EPIC Physical Activity Questionnaire
EPIC: European Prospective Investigation into Cancer
EPR: Electronic Patient Record
ER: Oestrogen Receptor
FFQ: food-frequency questionnaire
GCP: Good Clinical Practice
GDPR: General Data Protection Regulation
HRA: Health Research Authority
ICF: Informed Consent Form
ISRCTN: International Standard Randomised Controlled Trial Number
LTHT: Leeds Teaching Hospitals NHS Trust
MRI: Magnetic Resonance Imaging
NACT: Neoadjuvant Chemotherapy
NHS: National Health Service
PIS: patient information sheet
PR: Progesterone Receptor
PROMs: Patient Reported Outcome Measures
REC: Research Ethics Committee
SAE: Serious Adverse Event
TNBC: Triple Negative Breast Cancer
WCRF: World Cancer Research Fund

## Declarations

### Ethics approval and consent to participate

Ethical approval for the PRE-NUTRITIVE Study was granted 11^th^ of May 2022 by the West Midlands - Solihull Research Ethics Committee, reference: 22/WM/0087. Current protocol version 1.4 (4^th^ of April 2024). This trial was prospectively registered at ISRCTN on 12^th^ December 2022 (ISRCTN20130557) available at doi.org/10.1186/ISRCTN20130557).

### Consent for publication

Not applicable.

### Availability of data and materials

Not applicable.

### Authors’ contributions

JLT conceptualized the project; JLT obtained funding for the project; JLT, GC, BK, and SH developed the methodology and study design; GC wrote the original manuscript draft; JLT, GC, BK, SH, SDH, IB, AB, AR, NS, ERJV, TAH reviewed and edited the manuscript; JLT, GC, and BK acquired funding for the project; JLT, GC, and BK supervised the project; JLT and GC administered the project; GC, JLT, BK, SH designed the PRE-NUTRITIVE Study protocol; GC and JLT wrote the ethical approval application. All authors approved the final version.

## Acknowledgements

n/a

